# Underutilization of Syndrome-Specific ICD-10 Codes for Genetic Epilepsies: Implications for Precision Medicine

**DOI:** 10.1101/2025.09.12.25335652

**Authors:** Émile Moura Coelho da Silva, Tobias Brünger, Gary Taylor, Mousumi Sinha, Alison Merket, Anu Cherukara, Sunanjay Bajaj, Jessica Clark, Ludovica Montanucci, Emily A. Huth, Mariana Fauteux, Samden D. Lhatoo, Christian M. Boßelmann, Costin Leu, Rahil A. Tai, Dennis Lal

## Abstract

**Objective:** Syndrome-specific ICD-10 codes have the global potential to enhance patient identification for precision therapies, clinical trials, and research. However, their real-world uptake remains poorly understood. Thus, this study evaluated the utilization of syndrome-specific ICD-10 codes for monogenic epilepsies at a large academic medical center.

**Methods:** We queried an institutional genetic testing database to identify patients with pathogenic or likely pathogenic variants in ten epilepsy genes with established syndrome-specific ICD-10 codes *(CDKL5, EHMT1, KCNQ2, MECP2, MED13L, SCN1A, SHANK3, SLC13A5, SLC2A1, SYNGAP1*). Clinical encounters were extracted from the electronic health record (EHR), and patients were included if they had at least one encounter after the later of two dates: the implementation of the syndrome-specific code or the date of their genetic test result. Variants of uncertain significance were manually curated, and phenotypes for Rett and Dravet syndromes were reviewed to ensure accurate grouping.

**Results:** Of 83 patients with qualifying variants, 39 met all inclusion criteria. Despite confirmed diagnoses, only 22 of 39 (56.4%) patients were ever documented with a syndrome-specific ICD-10 code. Additionally, these codes were only utilized in 31.1% of all encounters and represented just 14.5% (235/1,626) of codes used. Uptake varied by syndrome, provider specialty, and encounter type, and increased over time. In the Dravet syndrome subgroup (N=23), generic epilepsy codes were documented in more than twice as many encounters as the Dravet-specific code (G40.83). When G40.83 was documented, other epilepsy codes were utilized less frequently, suggesting that clinicians may treat G40.83 as a substitute for broader epilepsy ICD-10 codes.

**Significance:** Syndrome-specific ICD-10 codes for monogenic epilepsies are underutilized and inconsistently applied, limiting their ability to support precision medicine, and research. Automated and patient-driven coding support, as well as integration of structured genetic data in the EHR, are needed to close the gap between code availability and clinical practice.

**Key Points:**

- Syndrome-specific ICD-10 codes for monogenic epilepsies are available but remain underutilized in clinical practice.
- Fewer than two-thirds of patients with molecular diagnoses were ever assigned their syndrome-specific ICD-10 code, and usage was inconsistent across encounters.
- Documentation of syndrome-specific ICD-10 codes varied by syndrome, provider specialty, and encounter type.
- In Dravet syndrome, generic epilepsy codes were documented more than twice as often as the Dravet-specific code (G40.83). When G40.83 was utilized, other epilepsy codes were documented less frequently.
- Underutilization of syndrome-specific codes may limit patient identification for precision therapies, clinical trials, and rare disease research.

## 1 Introduction

Advances in genomic technologies, particularly next-generation sequencing, have transformed the diagnostic landscape of epilepsies, enabling rapid identification of genetic etiologies and the discovery of over 1,000 epilepsy-associated genes^1,2^. This growing understanding of epilepsy genetics has paved the way for precision medicine, in which management and treatment strategies are guided by an individual’s specific genetic etiology. Many gene-specific interventions are already in practice or in late-phase clinical trials, including the ketogenic diet for GLUT1 deficiency syndrome, ganaxolone for cyclin-dependent kinase-like 5 (*CDKL5*) deficiency disorder, and antisense oligonucleotide therapy for *SCN1A*-related Dravet syndrome^3–5^. These advances underscore the importance of accurately making and documenting etiology-based diagnoses. As the number of genetic epilepsies and targeted treatments continues to increase, scalable methods to identify affected individuals within healthcare systems are urgently needed.

The International Classification of Diseases, Tenth Revision (ICD-10), is a standardized system of diagnosis codes developed by the World Health Organization and used internationally to record, report, and monitor diseases, enabling global comparisons of health data^6^. Utilization of appropriate ICD codes in the electronic health record (EHR) enables consistent clinical documentation and supports data-driven care. However, this is complicated by a lack of specific ICD-10 codes for most rare diseases, including genetic epilepsies^7^. In addition, multiple barriers to genetic testing access likely leave many individuals with genetic epilepsies, particularly adults, undiagnosed^8,9^. As a result, patients are often documented with nonspecific codes, such as G40.8 (“Other epilepsy and recurrent seizures”), which fail to capture the full spectrum of their condition. To address this gap, revisions to the ICD system are periodically made to introduce new, more exact codes. While this a big step forward, it does not come without challenges, as securing a new ICD code for rare diseases is a time- and resource-intensive process, requiring collaboration among diverse stakeholders, including patient advocacy organizations, clinical experts, and researchers^10^. Despite this, several genetic epilepsies, such as Dravet syndrome (G40.83) and *KCNQ2*-related epilepsy (G40.84), have recently been assigned their own ICD-10 codes, and additional efforts are ongoing (Supplementary Figure 1)^11,12^. For advocacy groups, investing time and resources in the development of syndrome-specific ICD codes is crucial. Such codes are expected to improve patient identification, thereby streamlining access to targeted therapies, facilitating clinical trial enrollment, and enhancing research participation^11,12^. In contrast to genetic data, which is often stored in unstructured formats in the EHR such as scanned reports or free-text notes, ICD codes provide a more standardized and structured approach for patient identification^13^.

While specific ICD codes for rare diseases are becoming more common and are seen as a potential solution to many challenges in rare disease research and care, the mere existence of an ICD code does not guarantee its use. A 2018 study examining physician coding accuracy and completeness across standardized clinical scenarios found that only 56% of documented ICD-10 codes were rated as appropriate, while 26% of relevant codes were omitted altogether^14^. Another example of the disconnect between code creation and uptake can be observed in the case of Friedreich’s ataxia. When ICD-10 was introduced in 2015, Friedreich’s ataxia lost its unique ICD-9 code (334.0) and was grouped under a broader code for “Early-onset cerebellar ataxia” (G11.1), which includes multiple conditions, such as Machado-Joseph disease, making the diagnosis inaccurate^10,15^. Of the 1,276 patients who had been correctly identified under the ICD-9 code, only 558 (44%) were subsequently documented with the new ICD-10 code^10^. Overall, several barriers, including limited clinician awareness, evolving code definitions, suboptimal EHR search and auto-suggest workflows, and competing documentation priorities, continue to limit the consistent use of specific ICD codes for rare diseases^16–19^.

The identification of patients with genetic epilepsies remains a challenge in large electronic health systems, one that must be overcome to optimize precision care and clinical trial enrollment for disease-modifying therapies. Therefore, understanding how syndrome-specific ICD codes are utilized is essential. To the best of our knowledge, no systematic study to date has investigated the adoption of these codes in monogenic epilepsies. To address this gap, we evaluated the use of syndrome-specific ICD-10 codes for monogenic epilepsies at a large academic medical center by integrating genetic testing results with longitudinal clinical encounter data. This approach allowed us to assess how consistently these codes are applied in practice and to identify factors influencing their utilization. Our findings highlight that even when syndrome-specific codes are available, their underutilization may limit the ability to identify eligible patients for research, clinical trials, and precision therapies.

## 2 Materials and Methods

### 2.1 Selection of syndrome-specific ICD-10 codes

We obtained a list of 17 ICD-10 codes for rare epilepsies from the Rare Epilepsy Network in June 2025 (Supplementary Figure 1)^20^. Because our study focused on monogenic epilepsies, we excluded codes corresponding to non-genetic epilepsies (e.g., hypoxic-ischemic encephalopathy) and genetically heterogeneous syndromes without a single causative gene (e.g., Lennox-Gastaut syndrome). We also excluded codes for disorders driven primarily by non-single nucleotide genetic variants (SNV), such as ring chromosome 20, since our institutional genetic testing results database currently only captures variants affecting individual genes. After applying these filtering criteria, ten monogenic epilepsy syndromes with unique ICD-10 codes were retained (Table 1).

**TABLE 1.** List of monogenic epilepsies with a syndrome-specific ICD-10 code, associated genes, and code implementation dates.

| Syndrome | ICD-10 Code | Gene | Effective Date<sup>21</sup> |
| --- | --- | --- | --- |
| Rett syndrome | F84.2 | *MECP2* | 10/01/2015 |
| Glucose transporter protein type 1 deficiency syndrome (GLUT1-DS) | E74.810 | *SLC2A1* | 10/01/2020 |
| Cyclin-dependent kinase-like 5 deficiency disorder (CDD) | G40.42 | *CDKL5* | 10/01/2020 |
| Dravet syndrome | G40.83 | *SCN1A* | 10/01/2020 |
| SYNGAP1-related intellectual disability (SYNGAP1-ID) | F78.A1 | *SYNGAP1* | 10/01/2021 |
| MED13L syndrome | Q87.85 | *MED13L* | 10/01/2023 |
| Phelan-McDermid syndrome | Q93.52 | *SHANK3* | 10/01/2023 |
| SLC13A5 citrate transporter disorder | E74.820 | *SLC13A5* | 10/01/2024 |
| KCNQ2-related epilepsy | G40.84 | *KCNQ2* | 10/01/2024 |
| Kleefstra syndrome | Q87.86 | *EHMT1* | 10/01/2024 |

### 2.2 Cohort identification

The study cohort was identified at UT Physicians, the clinical practice of UTHealth Houston, a large academic center that provides specialized care across multiple specialties. UT Physicians is also a Level 4 Adult and Pediatric Epilepsy Center accredited by the National Association of Epilepsy Centers (NAEC). This retrospective study was approved by the Institutional Review Board at UTHealth Houston (HSC-MS-23-1129). The requirement for informed consent was waived as the study involved analysis of aggregate data with no identifiable patient information.

We queried our institutional genetic testing database in January 2025. This curated database contains clinical genetic testing results for 1,602 patients and integrates data from the EHR, genetic testing company portals, and clinician-maintained records. We identified 166 patients carrying a total of 173 variants in one of the ten monogenic epilepsy genes with syndrome-specific ICD-10 codes listed in Table 1 and available at the time of the study (Figure 1).

**Figure 1.**
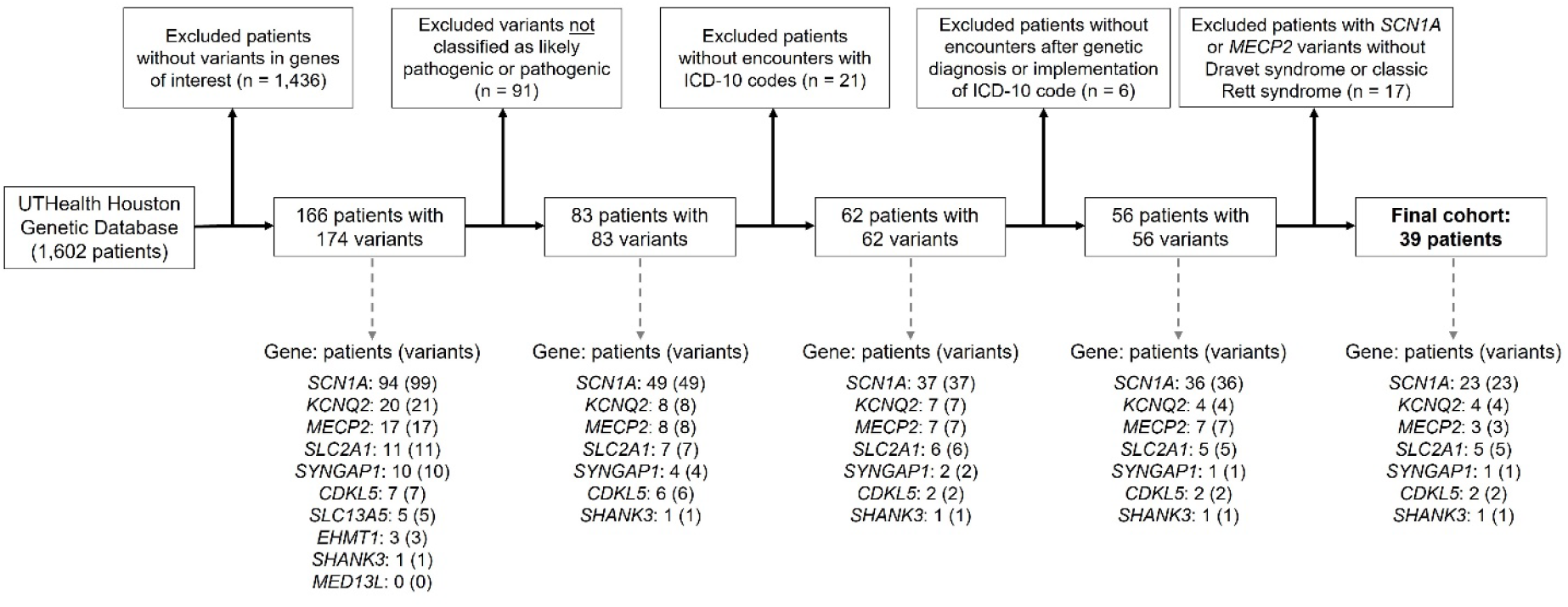
Workflow for patient cohort selection. Patients were selected from the UTHealth Houston genetic testing database based on variants in ten genes (listed in Table 1) associated with monogenic epilepsy syndromes. The figure outlines the filtering steps and final cohort composition.

We restricted our analysis to individuals with single nucleotide variants (SNVs) and intragenic copy-number variants classified as likely pathogenic or pathogenic, identifying 83 individuals (83/166, 50%). Patients with variants classified by the laboratory as likely benign or benign were excluded. Variants of uncertain significance (VUS) were annotated using an internally developed bioinformatic pipeline that incorporates data from the latest population and patient databases, as well as *in silico* prediction tools. This pipeline also applied the variant classification criteria established by the American College of Medical Genetics and Genomics and the Association for Molecular Pathology (ACMG/AMP), as well as recently published refinements of the guidelines^22,23^. All reclassifications were reviewed by a US certified genetic counselor and variant scientists specialized in epilepsy genes. Two VUSs reclassified as likely pathogenic or pathogenic were included in the analysis, while remaining VUSs were excluded.

For the 83 patients with a likely pathogenic or pathogenic variant, we extracted clinical data from the EHR in February 2025, including demographics, ICD-10 codes documented in each encounter, date of service, provider name and specialty, and encounter type (e.g., refill, office visit). Unique encounters were defined as those with a distinct combination of provider name and date of service. Only patients with encounters documented with at least one ICD-10 code were included, yielding 62 patients (62/83, 74.7%). Additionally, we only included patients that had encounters on or after whichever date came later: (1) the implementation date of the syndrome-specific ICD-10 code or (2) the date the genetic testing report was issued. This resulted in a total of 56 patients (56/62, 77.8%).

Finally, because variants in the *MECP2* and *SCN1A* genes can cause multiple phenotypes but the ICD-10 codes are specific to Rett and Dravet syndromes, we reviewed clinical phenotypes to confirm consistency with the diagnoses of interest. Individuals with *MECP2* variants were included only if they met clinical criteria for classic Rett syndrome^24^. Similarly, only patients with *SCN1A* variants whose clinical presentation was consistent with Dravet syndrome were retained^25^. This led to the exclusion of thirteen patients with *SCN1A* variants and four patients with *MECP2* variants, resulting in a final cohort of 39 patients (Supplementary Table 1).

### 2.3 Statistical analysis

Statistical analyses were performed using R, version 4.4.0^26^. Continuous variables (e.g., patient age, number of encounters) are reported as median with interquartile range (IQR) and compared between groups using the Wilcoxon rank-sum test. Statistical significance was defined as p<0.05 and visualizations were generated with ggplot2.

## 3 Results

In this study, we evaluated the real-world use of syndrome-specific ICD-10 codes for monogenic epilepsies. By analyzing ten established codes, we quantified the proportion of eligible patients who were documented with the syndrome-specific code and identified factors influencing coding uptake. Bridging this gap between code availability and use is critical, as underutilization can limit access to precision therapies, clinical trials, and research opportunities.

### 3.1 Clinical and genetic characteristics of the study cohort

To identify patients eligible for the ten syndrome-specific ICD-10 codes, we queried our institutional genetic testing database for individuals with likely pathogenic or pathogenic variants in the corresponding genes and extracted their clinical data from the EHR (see Materials and Methods for details; Figure 1). For three genes with syndrome-specific ICD-10 codes, *EHMT1, MED13L*, and *SLC13A5*, no patients with likely pathogenic or pathogenic variants were identified. As a result, the final cohort represents seven monogenic epilepsies. Two *SCN1A* variants initially classified as variants of uncertain significance were reclassified as likely pathogenic after expert review and were therefore included in the final cohort.

After excluding individuals without encounters containing at least one ICD-10 code or without a matching clinical phenotype, the final cohort consisted of 39 patients: 23 with Dravet syndrome, five with GLUT1 deficiency syndrome (GLUT1-DS), four with KCNQ2-related epilepsy, three with Rett syndrome, two with *CDKL5* deficiency disorder (CDD), one with SYNGAP1-related intellectual disability (SYNGAP1-ID), and one with Phelan-McDermid syndrome. Although not the primary focus of our study and thus excluded from downstream analyses, we identified six individuals documented with syndrome-specific ICD-10 codes without confirmed genetic diagnoses, suggesting possible misclassification (see Supplemental Table 2).

The final cohort of 39 patients had a median age of 9.7 years (IQR: 5.3–14.5 years), and a total of 1,122 clinical encounters since their genetic diagnoses and the release of the relevant syndrome-specific ICD-10 codes. Across these encounters, clinicians documented 2,433 ICD-10 diagnosis codes. Despite all 39 patients being eligible for a syndrome-specific ICD-10 code based on their genetic diagnosis and clinical presentation, only 22 (56.4%) patients were ever documented with one. The remaining 17 (43.6%) patients were coded exclusively with more general diagnosis codes, such as “unspecified epilepsy” (ICD-10: G40.9) or “genetic susceptibility to disease” (ICD-10: Z15). While patients documented with a syndrome-specific ICD-10 code at least once had a higher number of encounters (median: 27, IQR: 8.0–42.5) compared to those without such a code (median: 4.5, IQR: 1–36.3), this difference was not statistically significant (p=0.057, Wilcoxon rank-sum test).

### 3.2 Differences in syndrome-specific ICD-10 code use by syndrome, time, provider specialty, and encounter type

The utilization of syndrome-specific ICD-10 codes was inconsistent across the different syndromes. All four patients with GLUT1-DS and all three patients with Rett syndrome had E74.810 and F84.2 documented in their charts at least once, respectively (Figure 2A). In contrast, only 14 of 23 (60.9%) patients with Dravet syndrome had G40.83 documented. The one patient with SYNGAP1-ID had F78.A1 documented once, while none of the patients with *CDD* or KCNQ2-realted epilepsy patients had G40.42 or G40.84 in their charts, respectively. Inconsistency in the utilization of syndrome-specific ICD-10 codes was also evident at the encounter level. While these codes were documented in nearly half (20/42, 47.6%) of encounters for patients with Rett syndrome and 30.9% (17/55) of encounters for patients with GLUT1-DS (F84.2 and E74.810, respectively), documentation was markedly lower for other syndromes—23.1% (196/849) for Dravet syndrome and 12.5% (1/8) for SYNGAP1-ID (Figure 2B).

**Figure 2.**
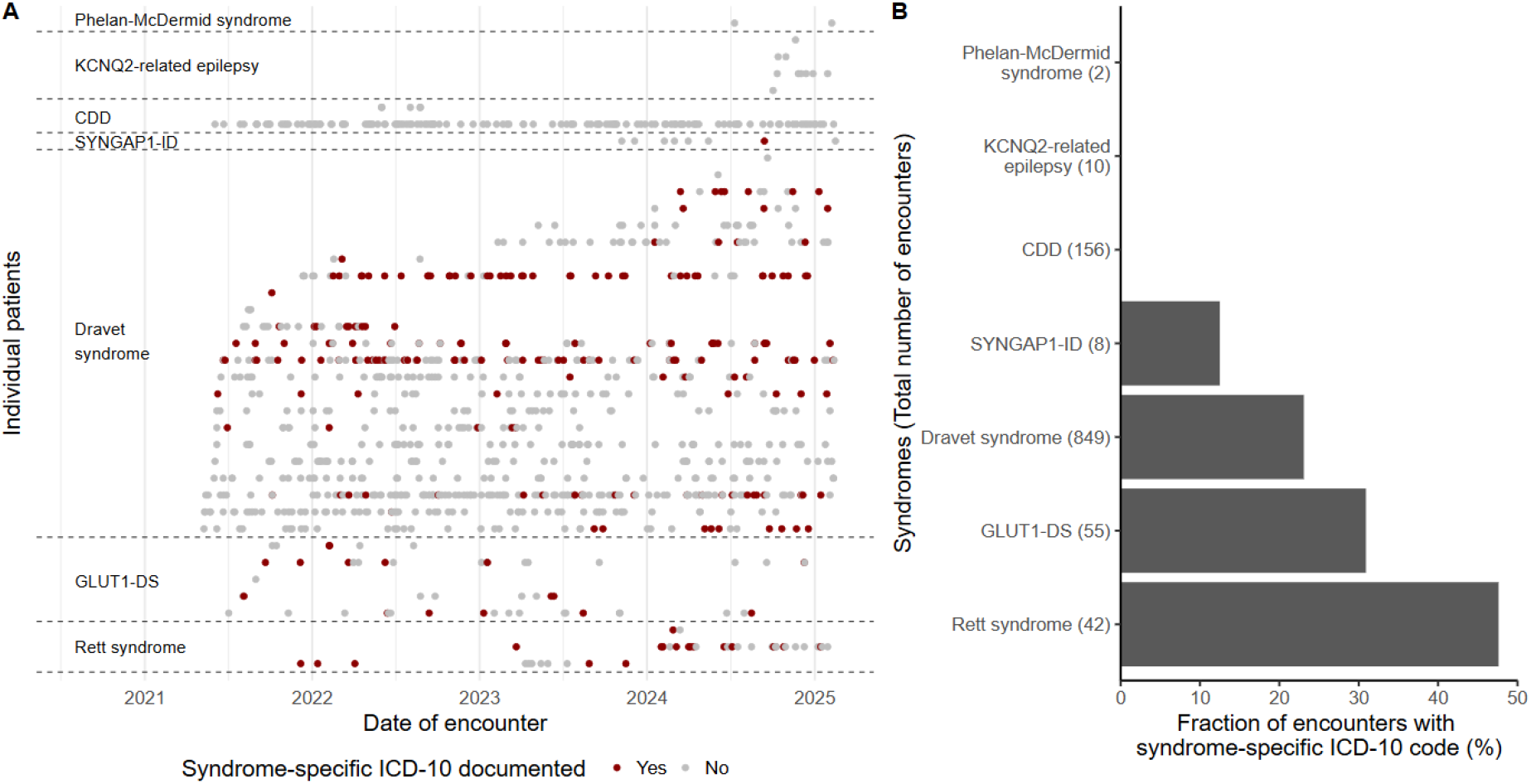
Utilization of syndrome-specific ICD-10 codes across clinical encounters. Clinical encounters for individuals with molecularly confirmed genetic epilepsies are presented only for the period following the implementation of each gene-specific ICD-10 code. (A) Longitudinal utilization of syndrome-specific ICD-10 code by gene. Each horizontal line represents an individual patient, and each dot represents a clinical encounter. Red dots indicate encounters where the syndrome-specific ICD-10 code was documented; gray dots indicate encounters without such coding. (B) Proportion of encounters in which the syndrome-specific ICD-10 code was documented, stratified by syndrome. Numbers in parentheses indicate the total number of encounters for patients with that syndrome. Abbreviations: CDD: cyclin-dependent kinase-like 5 (CDKL5) deficiency disorder; SYNGAP1-ID: SYNGAP1-related intellectual disability; GLUT1-DS: glucose transporter protein type 1(GLUT1) deficiency syndrome.

To further investigate these inconsistencies, we examined the subset of 22 patients who had at least one encounter documented with a syndrome-specific ICD-10 code. Across their 752 encounters, only 234 (31.1%) had a syndrome-specific code (Supplementary Figure 2A). Notably, this underutilization was not due to a lack of coding overall. Each encounter could have multiple ICD-10 codes, and across the 752 encounters, a total of 1,626 codes were documented, of which only 235 (14.5%) were syndrome-specific (Supplementary Figure 2B). Moreover, when a syndrome-specific code was documented in one visit, it was often omitted in subsequent encounters, even when those visits were with the same specialty or provider (Figure 2A). Despite this sporadic use, documentation of syndrome-specific ICD-10 codes increased over time, from 13.2% of encounters (19/144) in 2021 to 28.2% (86/305) in 2024 (Supplementary Figure 3A). This upward trend was also observed in the Dravet syndrome subgroup and when analyses were restricted to neurology encounters (Supplementary Figures 3B–3D).

Next, we explored if provider specialty or encounter type influenced the use of syndrome-specific ICD-10 codes, and both factors affected code utilization. Specialties such as Pediatric Pulmonology documented these codes in 50.0% (11/22) of encounters, whereas specialties such as Pediatric Neurology and Neurology documented them in 26.3% (152/577) and 9.3% (14/150) of encounters, respectively (Figure 3A). Similarly, syndrome-specific ICD-10 codes were documented in 49.2% (31/63) of telemedicine encounters but only used in 15.4% (2/13) of ancillary procedure encounters and 10.0% (41/411) of refill encounters (Figure 3B).

**Figure 3.**
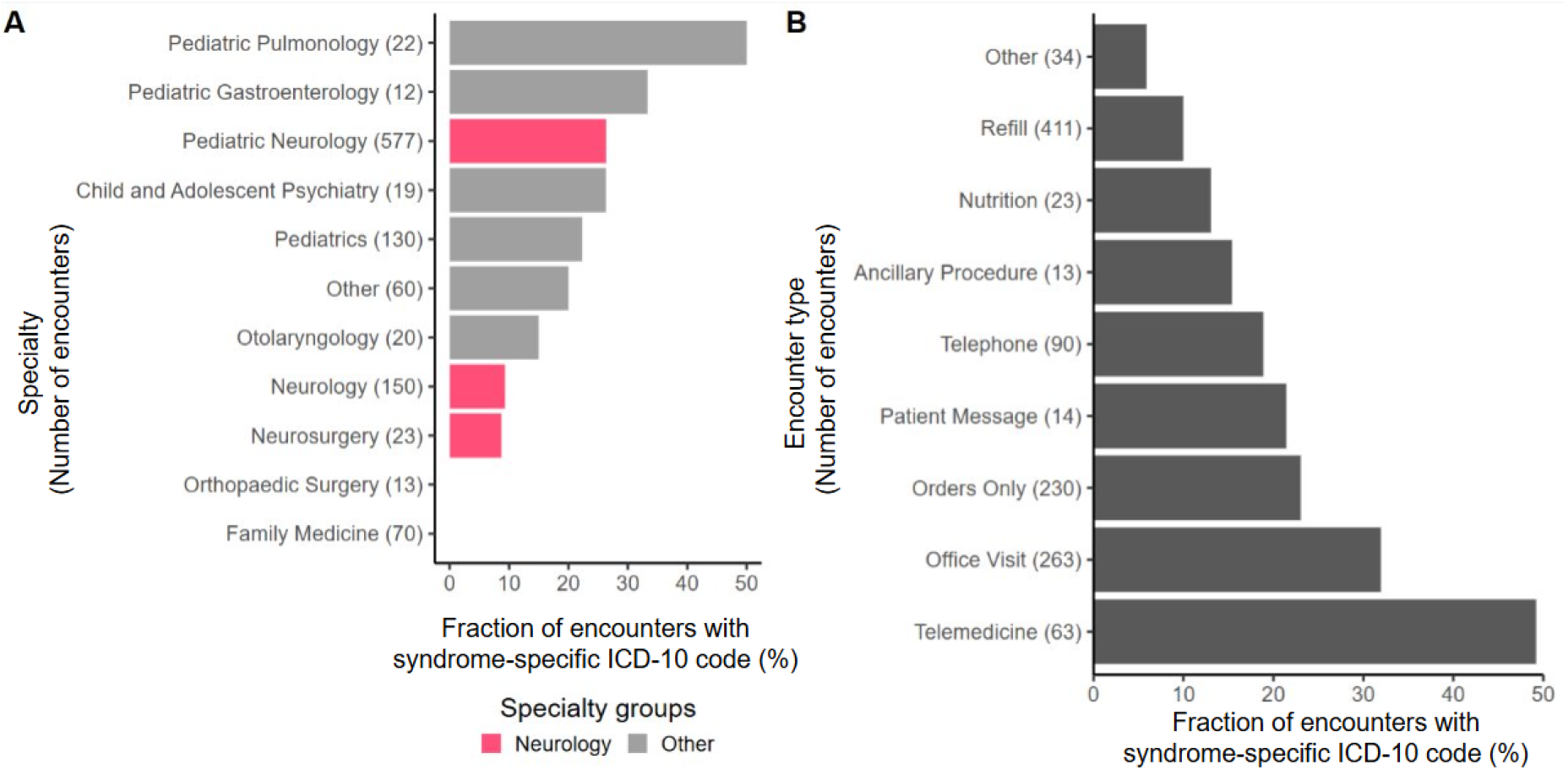
Utilization of syndrome-specific ICD-10 codes by provider specialty and encounter type. (A) Fraction of encounters in which a syndrome-specific ICD-10 code was documented, grouped by provider specialty. (B) Fraction of encounters in which a syndrome-specific ICD-10 code was documented, grouped by encounter type. Specialties and encounters with fewer than 10 counts were grouped under “Other.” “Telemedicine” refers to a full virtual visit, whereas “Telephone” indicates brief calls, typically for results disclosure or other follow-up. Plot A “Other”: Cardiology, Clinical Genetics and Genomics, Dermatology, General Surgery, Lab, Ophthalmology, Pediatric Cardiology, Pediatric Endocrinology, Pediatric Hematology, Pediatric Neurosurgery, Pediatric Surgery, Pediatric Urology, Physical Medicine and Rehabilitation, Plastic Surgery, Psychiatry, Psychology, Pulmonary Disease, Social Services. Plot B “Other”: Ancillary Orders, Ancillary Orders, Appointment, Consult, Documentation, Immunization, Nurse Only, Patient Outreach, Procedure Visit, Social Work, Transcribe Orders, Transcribe Orders.

### 3.3 Syndrome-specific ICD-10 code replaces generic epilepsy codes in Dravet syndrome subcohort

Given that Dravet syndrome was the largest subgroup in our cohort (23 patients), we examined coding patterns in greater detail, including patients both with and without the syndrome-specific ICD-10 code. This subgroup was likely the largest because Dravet syndrome can often be clinically diagnosed and is strongly associated with a single gene, unlike most other monogenic epilepsies. Across 849 encounters, the majority (635/849, 76.0%) were documented with at least one epilepsy-related ICD-10 code (G40) (Figure 4). Generic epilepsy codes were used more than twice as often (439/635, 69.1%) as the Dravet-specific code (196/635, 30.9%). When the Dravet code (G40.83) was documented, other epilepsy codes were used less frequently (Supplementary Tables 3– 4), suggesting that clinicians may treat G40.83 as a substitute for broader epilepsy ICD-10 codes. We also explored other diagnosis codes frequently documented in this subcohort. Notably, every encounter coded with G40.83 also included Z15.1 (Genetic susceptibility to epilepsy and neurodevelopmental disorders), suggesting a local EHR configuration that auto-populates this code when G40.83 is selected (Supplementary Table 3).

**Figure 4.**
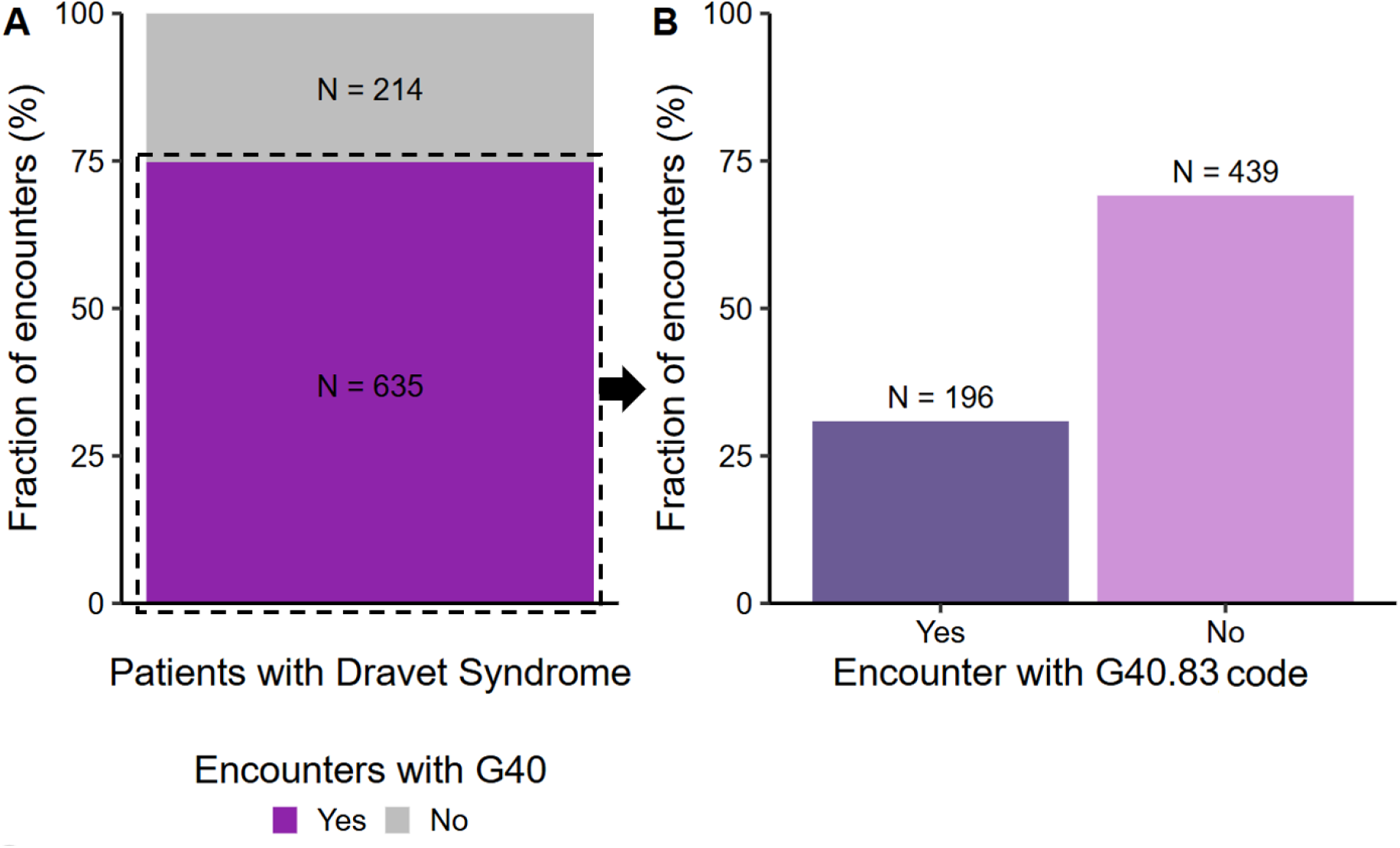
Epilepsy ICD-10 utilization in patients with Dravet syndrome. (A) Fraction of encounters with that included an “epilepsy and recurrent seizures” ICD-10 code (G40). (B) Breakdown of G40-coded encounters by use of Dravet syndrome code (G40.83) versus other G40 codes.

## 4 Discussion

ICD codes are globally important as they provide a standardized framework for recording and comparing diseases across countries, supporting clinical care, research, reimbursement, health policy, and public health reporting. In this first assessment of the utilization of syndrome-specific ICD-10 codes for monogenic epilepsies, we demonstrate a significant underutilization: only 56.4% of eligible patients had their syndrome-specific code documented in their charts at least once. The remainder of the cohort was documented exclusively with nonspecific codes, which fail to reflect the distinct clinical and genetic features of the disorders (e.g., epilepsy, movement disorders) and limit the ability to differentiate these patients from broader epilepsy populations. Even in patients who were documented with a syndrome-specific code at least once, these codes accounted for only 31.1% of their total encounters and just 14.5% of all codes used overall. Moreover, their application was inconsistent, with later encounters frequently reverting to nonspecific coding. Such inconsistency may impact continuity of care and further complicate efforts to track outcomes over time. We also observed that utilization of syndrome-specific codes varied by syndrome, time, and clinical context (provider specialty and encounter type), suggesting that certain settings or conditions foster better uptake than others. Overall, this underutilization of available syndrome-specific ICD codes impedes the efficient identification of patients for research, clinical trials, and targeted treatments.

As expected, our findings show that the establishment of a dedicated ICD code does not ensure its consistent use in clinical practice. Prior studies in other clinical contexts have similarly demonstrated the underutilization of diagnosis codes, highlighting their low sensitivity for case identification^27,28^. For example, in an obstetric population, the sensitivity of ICD codes to capture pregnancy complications ranged from 0.15 to 0.75^27^. In this context, our observation that 43.6% of genetically confirmed epilepsy patients lacked documentation of their syndrome-specific code is consistent with these patterns of incomplete coding uptake. Although some specialties used syndrome-specific ICD-10 codes more frequently, the overall underutilization and inconsistent documentation across specialties highlight persistent gaps in awareness and coding practices. Encouragingly, we observed a gradual increase in coding rates over time within our cohort, suggesting that uptake of syndrome-specific codes improves with time and growing provider awareness. One possible contributor is the role of patient advocacy organizations, which actively promote the use of newly established ICD codes and encourage families to share them with their providers, efforts that may accelerate dissemination and adoption^11,29^.

Another notable finding in our study was the variation in code documentation by syndrome. Some syndrome-specific ICD-10 codes, such as G40.84 for KCNQ2-related epilepsy, were never used, likely because this code only became available in October 2024, leaving little time for providers to incorporate it into practice. In contrast, other syndrome-specific codes, such as G40.42 for CDKL5 deficiency disorder, have been available for many years yet were never utilized. This slow adoption stands in contrast to other conditions, such as *Clostridioides difficile* infection, where updated ICD-10 codes were adopted immediately without latency^30^. This suggests that additional factors, such as EHR workflows, reimbursement practices, or coding habits may contribute to inconsistent uptake. Previous studies have identified several barriers to accurate and thorough coding, including limited knowledge of ICD codes, constraints on documentation time, and tendency to carry forward diagnosis codes from prior encounters^16,18,19^. These challenges are likely amplified in the context of rare diseases, where diagnosis codes are often lacking or overly broad, and genetic testing reports are frequently unstructured and difficult to locate in the EHR^7,13,31^.

Our study provides the first systematic assessment of real-world utilization of syndrome-specific ICD-10 codes for monogenic epilepsies. However, several limitations should be acknowledged. This was a retrospective analysis conducted at UTHealth Houston (UT Physicians), a large academic center that provides specialized care across multiple specialties. The study included a relatively small sample size of 39 patients from a single center, which may limit the generalizability of our findings. In particular, the observed coding practices may not reflect those in community hospitals, rural settings, or other healthcare systems with different resources and workflows. Additionally, some syndrome-specific ICD-10 codes were only recently implemented, leaving limited time for adoption by providers. Nonetheless, our findings reveal a clear gap between the availability of syndrome-specific ICD-10 codes for monogenic epilepsies and their clinical use. This underutilization of syndrome-specific ICD-10 codes reflects broader barriers to genetic-informed care, including limited clinician training in genetics and inconsistent access to genetic experts, such as genetic counselors^8,32^.

Moving forward, interventions to support clinicians and medical coders in the assignment of ICD codes are essential to improve the utilization of syndrome-specific ICD-10 codes. AI-assisted coding, employing techniques such as natural language processing, can analyze free-text clinical notes alongside structured EHR data to generate real-time diagnosis code suggestions. This approach enables clinicians to review and refine proposed codes rather than coding entirely manually. Previous studies evaluating similar methods have shown promise in improving coding efficiency and, in some cases, accuracy^33–36^. Patient-driven coding support, in which patients select possible symptoms or conditions that correspond to specific ICD codes on intake forms, could also offer a complementary strategy. Additionally, as EHR systems such as Epic evolve to support structured genetic data, providers may also more easily access patients’ genetic diagnoses, which could in turn increase the likelihood of using corresponding syndrome-specific codes^37^. System-level changes may also play a role; for example, the transition to ICD-11, available for global use since 2022, expands coding to over 5,500 rare diseases, potentially improving their visibility in the medical record^38^. Nonetheless, relying solely on ICD codes for patient identification is insufficient. Complementary approaches, such as the use of rare disease registries, should be employed concurrently to maximize identification of eligible individuals for targeted therapies, clinical trials, and research studies^39,40^. Finally, it is important to recognize that even when ICD codes are applied, they cannot fully capture the underlying heterogeneity of genetic conditions, where variants with different functional effects may lead to distinct phenotypes and require different therapeutic approaches^41,42^.

Future research should explore syndrome-specific ICD-10 code utilization across multiple institutions to determine whether similar underutilization occurs. It will also be important to assess factors contributing to underutilization, such as individual provider or medical coder practices or EHR system features.

## 5 Conclusion

Syndrome-specific ICD-10 codes for monogenic epilepsies are markedly underutilized, even for patients with confirmed molecular diagnoses and established clinical syndromes. In our cohort, fewer than two-thirds of eligible patients were ever documented with their syndrome-specific ICD-10 code, and when used, these codes were applied inconsistently across encounters, specialties, and time. Such gaps hinder the reliable identification of patients for precision therapies, clinical trials, and research studies, limiting the intended value of these codes. Although uptake of syndrome-specific ICD-10 codes showed gradual improvement over time, additional efforts, including automated and patient-driven coding support and integration of structured genetic data, are needed to ensure accurate and consistent use. Broader, multi-institutional studies will be essential to validate these findings and to guide strategies that maximize the clinical and research utility of syndrome-specific ICD codes as precision medicine advances.

## Data Availability

All data produced in the present study are available upon reasonable request to the authors.

## Author Contributions

Émile Moura Coelho da Silva: study concept and design, data acquisition, cohort identification, data analysis, data visualization, first draft of manuscript, and review and editing of manuscript. Tobias Brünger: study concept and design, data acquisition, cohort identification, data analysis, data visualization, and review and editing of manuscript. Gary Taylor: data acquisition, cohort identification, review and editing of manuscript. Mousumi Sinha: data acquisition, review and editing of manuscript. Alison Merket: cohort identification, review and editing of manuscript. Anu Cherukara: cohort identification, review and editing of manuscript. Sunanjay Bajaj: cohort identification, review and editing of the manuscript. Jessica Clark: data acquisition, data analysis, review and editing of manuscript. Ludovica Montanucci: cohort identification, review and editing of manuscript. Emily Huth: cohort identification, review and editing of manuscript. Mariana Fauteux: cohort identification, review and editing of manuscript. Samden Lhatoo: review and editing of manuscript. Christian M Boßelmann: review and editing of manuscript. Costin Leu: cohort identification, review and editing of manuscript. Rahil A. Tai: data acquisition, review and editing of manuscript. Dennis Lal: study concept and design, review and editing of manuscript, and project supervision. All authors read and approved the final manuscript.

## Acknowledgements

None.

## Data Availability Statement

All data produced in the present study are available upon reasonable request to the authors.

