## Supplemental Data for "Underutilization of Syndrome-Specific ICD-10 Codes for Genetic Epilepsies: Implications for Precision Medicine"

### Supplementary Figures

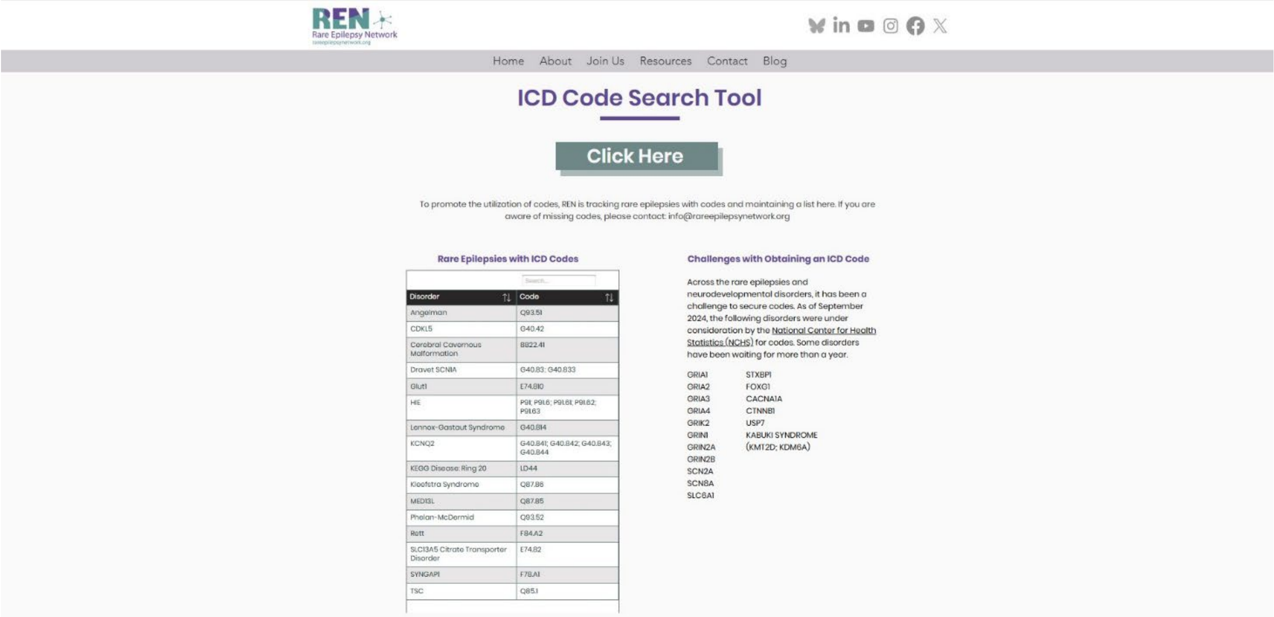


| Disorder | Code |
|---|---|
| Angelman | Q93.51 |
| CDKL5 | G40.42 |
| Cerebral Cavernous Malformation | B822.41 |
| Dravet SCN1A | G40.83; G40.833 |
| Glut1 | E74.810 |
| HIE | P91; P91.6; P91.61; P91.62; P91.63 |
| Lennox-Gastaut Syndrome | G40.814 |
| KCNQ2 | G40.841; G40.842; G40.843; G40.844 |
| KEGG Disease: Ring 20 | LD44 |
| Kleefstra Syndrome | Q87.86 |
| MED13L | Q87.85 |
| Phelan-McDermid | Q93.52 |
| Rett | F84.A2 |
| SLC13A5 Citrate Transporter Disorder | E74.82 |
| SYNGAP1 | F78.A1 |
| TSC | Q85.1 |

**Supplementary Figure 1. Rare epilepsies with approved ICD-10 codes (as of 06-30-2025).** Screenshot from the Rare Epilepsy Network website showing all rare epilepsies for which a syndrome-specific ICD-10 code has been established.

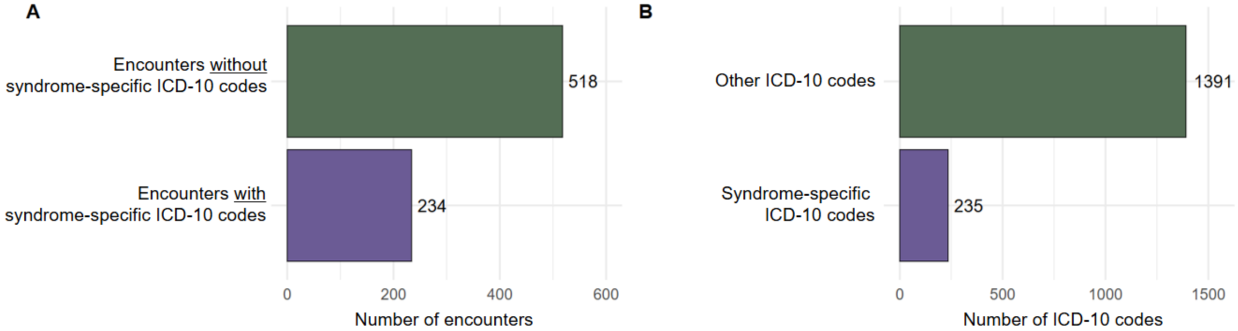


**Supplementary Figure 2. Total encounters and ICD-10 codes for patients with a documented syndrome-specific ICD-10 code (N=24 patients).**
***(A)*** *Total number of encounters, split by utilization of a syndrome-specific ICD-10 code.*
***(B)*** *Total ICD-codes, split by syndrome-specific ICD-10 codes and other ICD-10 codes.*

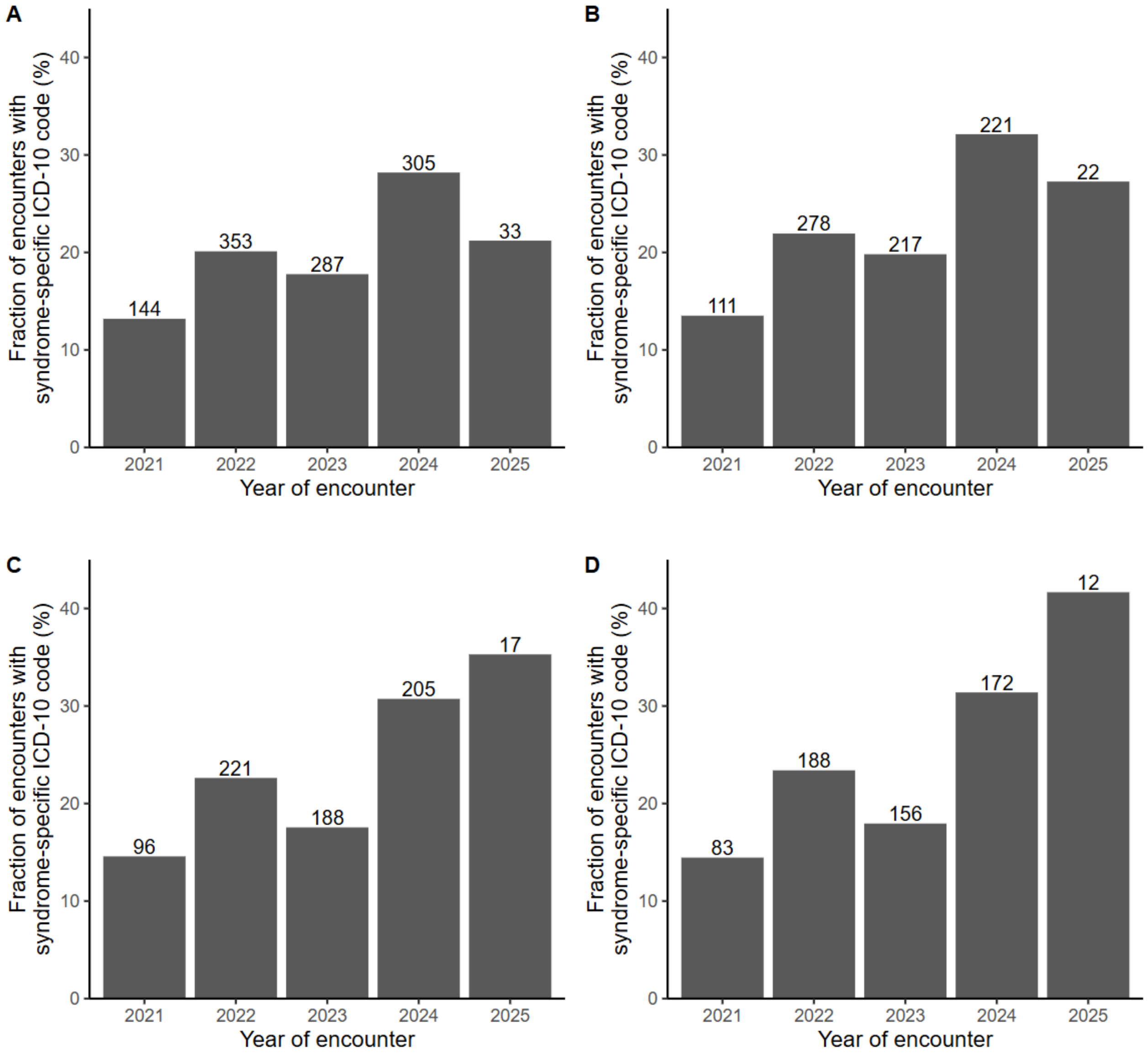


**Supplementary Figure 3. Utilization of syndrome-specific ICD-10 codes over time.** ***(A)*** *Proportion of encounters coded with a syndrome-specific ICD-10 code across all syndromes.* ***(B)*** *Same as (A), limited to patients with Dravet syndrome.* ***(C)*** *Proportion of neurology encounters coded with a syndrome-specific ICD-10 code across all syndromes.* ***(D)*** *Same as (C), limited to patients with Dravet syndrome.*

### Supplementary Tables

#### Supplementary Table 1. Genetic variants identified in the final cohort.

| ID | Gene | Transcript | Variant | Classification | Year of Genetic Testing Report |
|---|---|---|---|---|---|
| 1 | *CDKL5* | NM_003159 | c.1648C>T (p.Arg550*) | Pathogenic | 2020 |
| 2 | *CDKL5* | NM_003159 | c.2046del (p.Gly683Valfs*101) | Pathogenic | 2022 |
| 3 | *KCNQ2* | NM_172107 | c.2114dupC (p.Ala706Glyfs*159 | Likely pathogenic | 2019 |
| 4 | *KCNQ2* | NM_172107 | c.1273_1274del (p.Leu425Valfs*9) | Likely pathogenic | 2023 |
| 5 | *KCNQ2* | NM_172107 | Deletion exon 6-17 | Pathogenic | 2022 |
| 6 | *KCNQ2* | NM_172107 | c.1273_1274del (p.Leu425Valfs*9) | Likely pathogenic | 2023 |
| 7 | *MECP2* | NM_004992 | c.880C>T (p.Arg294*) | Pathogenic | 2023 |
| 8 | *MECP2* | NM_004992 | c.917G>T (p.Arg306Leu) | Pathogenic | 2021 |
| 9 | *MECP2* | NM_004992 | c.473C>T (p.Thr158Met) | Pathogenic | 2022 |
| 10 | *SCN1A* | NM_001165963 | c.1377G>A (p.Gln459=)$^{+}$ | Likely pathogenic | 2024 |
| 11 | *SCN1A* | NM_001165963 | c.3794T>C (p.Leu1265Pro) | Pathogenic | 2008 |
| 12 | *SCN1A* | NM_001165963 | c.1170+1G>C | Pathogenic | 2010 |
| 13 | *SCN1A* | NM_001165963 | c.2134C>T (p.Arg712*) | Pathogenic | 2012 |
| 14 | *SCN1A* | NM_001165963 | c.2758_2764del (p.Val920Argfs*13) | Pathogenic | 2011 |
| 15 | *SCN1A* | NM_001165963 | Deletion exon 2 | Pathogenic | 2011 |
| 16 | *SCN1A* | NM_001165963 | c.3794T>C (p.Leu1265Pro) | Likely pathogenic | 2014 |
| 17 | *SCN1A* | NM_001165963 | c.4547C>A (p.S1516*) | Pathogenic | 2013 |
| 18 | *SCN1A* | NM_001165963 | c.677C>T (p.Thr226Met) | Likely pathogenic | 2014 |
| 19 | *SCN1A* | NM_001165963 | c.4786C>T (p.Arg1596Cys) | Pathogenic | 2020 |
| 20 | *SCN1A* | NM_001165963 | c.384dupA (p.Leu129Ilefs*21) | Pathogenic | 2016 |
| 21 | *SCN1A* | NM_001165963 | c.2362G>A (p.Glu788Lys) | Likely pathogenic | 2016 |
| 22 | *SCN1A* | NM_001165963 | c.2799C>A (p.His933Gln) | Likely pathogenic | 2016 |
| 23 | *SCN1A* | NM_001165963 | c.4906C>T (p.R1636*) | Pathogenic | 2019 |
| 24 | *SCN1A* | NM_001165963 | c.677C>T (p.Thr226Met) | Pathogenic | 2018 |
| 25 | *SCN1A* | NM_001165963 | c.229dupC (p.Leu77Profs*11) | Pathogenic | 2019 |
| 26 | *SCN1A* | NM_001165963 | c.2856G>A (p.Trp952*) | Pathogenic | 2021 |
| 27 | *SCN1A* | NM_001165963 | c.4786C>T (p.Arg1596Cys) | Pathogenic | 2021 |
| 28 | *SCN1A* | NM_001165963 | c.1178G>A (p.Arg393His) | Pathogenic | 2023 |
| 29 | *SCN1A* | NM_001165963 | Deletion exons 22-23 | Pathogenic | 2023 |
| 30 | *SCN1A* | NM_001165963 | c.3733C>T (p.Arg1245*) | Pathogenic | 2021 |
| 31 | *SCN1A* | NM_001165963 | c.5361 G>C (p.Glu1787Asp) | Likely pathogenic | 2023 |
| 32 | *SCN1A* | NM_001165963 | c.1096G>C (p.Asp366His) | Likely pathogenic | 2011 |

| | | | | | |
|---|---|---|---|---|---|
| 33 | *SHANK3* | NM_033517 | c.1760 G>A (p.Arg587Gln) | Likely pathogenic | 2024 |
| 34 | *SLC2A1* | NM_006516 | c.798dupC (p.Ala267Argfs*114) | Likely pathogenic | <2016 |
| 35 | *SLC2A1* | NM_006516 | c.1158_1162delCCCAT (p.Ile386Metfs*6) | Pathogenic | 2016 |
| 36 | *SLC2A1* | NM_006516 | c.694C>T (p.Arg232Cys) | Pathogenic | 2021 |
| 37 | *SLC2A1* | NM_006516 | c.388G>A (p.Gly130Ser) | Pathogenic | 2020 |
| 38 | *SLC2A1* | NM_006516 | c.1257C>T (p.Gly419=)[+] | Pathogenic | 2021 |
| 39 | *SYNGAP1* | NM_006772 | c.928G>A (p.Glu310Lys) | Pathogenic | 2023 |

[+]Synonymous variants expected to affect splicing.

**Supplementary Table 2. Patients documented with syndrome-specific ICD-10 codes that did not meet study eligibility criteria.**

| ID | ICD-10 Code | Gene | Variant | Classification | Exclusion Criteria |
|---|---|---|---|---|---|
| 40 | F84.2 | *MECP2* | c.808C>T(p.R270*) | Pathogenic | Not classic Rett syndrome (no developmental regression) |
| 41 | G40.834 | *SCN1A* | c.4887C>T(p.Phe1629Phe) | Uncertain | Not Dravet syndrome, variant is uncertain |
| 42 | G40.834/G40.833 | *SCN1A* | c.5582G>A(p.R1861Q) | Uncertain | Not Dravet syndrome, variant is uncertain |
| 43 | F84.2 | *GABR2* | c.911C>T(p.A304V) | Likely pathogenic | Variant not in a gene associated with the ICD-10 code |
| 44 | E74.810 | *SLC2A1* | c.1096_1098delinsAA (p.Y366N) | Pathogenic | Code used prior to genetic testing report |

**Supplementary Table 3. Fifteen most frequent ICD-10 codes documented across 196 encounters <u>with</u> a Dravet-syndrome code (G40.83).**

| ICD-10 Code | Code description | Number of Encounters | Fraction of Encounters (%) |
|---|---|---|---|
| Z15.1 | Genetic susceptibility to epilepsy and neurodevelopmental disorders | 196 | 100 |
| G40.834 | Dravet syndrome, intractable, without status epilepticus | 131 | 66.8 |
| G40.833 | Dravet syndrome, intractable, with status epilepticus | 66 | 33.7 |
| G40.319 | Generalized idiopathic epilepsy and epileptic syndromes, intractable, without status epilepticus | 28 | 14.3 |
| Q99.8 | Other specified chromosome abnormalities | 20 | 10.2 |
| F84.0 | Autistic disorder | 16 | 8.2 |
| R56.9 | Unspecified convulsions | 14 | 7.1 |
| G47.33 | Obstructive sleep apnea | 12 | 6.1 |
| R62.50 | Unspecified lack of expected normal physiological development in childhood | 11 | 5.6 |
| F90.2 | Attention-deficit hyperactivity disorder, combined type | 10 | 5.1 |

| | | | |
|---|---|---|---|
| F80.9 | Developmental disorder of speech and language, unspecified | 8 | 4.1 |
| G40.419 | Other generalized epilepsy and epileptic syndromes, intractable, without status epilepticus | 8 | 4.1 |
| Q76.1 | Klippel-Feil syndrome | 7 | 3.6 |
| F06.30 | Mood disorder due to known physiological condition, unspecified | 6 | 3.1 |
| G40.909 | Epilepsy, unspecified, not intractable, without status epilepticus | 6 | 3.1 |

**Supplementary Table 4. Fifteen most frequent ICD-10 codes documented across 439 encounters with<u>out</u> a Dravet-syndrome code (G40.83).**

| ICD-10 Code | Code Description | Number of Encounters | Fraction of Encounters (%) |
|---|---|---|---|
| G40.319 | Generalized idiopathic epilepsy and epileptic syndromes, intractable, without status epilepticus | 143 | 32.6 |
| G40.219 | Localization-related (focal) (partial) symptomatic epilepsy and epileptic syndromes with complex partial seizures, intractable, without status epilepticus | 79 | 18.0 |
| G40.909 | Epilepsy, unspecified, not intractable, without status epilepticus | 55 | 12.6 |
| G40.419 | Other generalized epilepsy and epileptic syndromes, intractable, without status epilepticus | 47 | 10.7 |
| G40.814 | Lennox-Gastaut syndrome, intractable, without status epilepticus | 43 | 9.8 |
| G40.919 | Epilepsy, unspecified, intractable, without status epilepticus | 39 | 8.9 |
| G40.309 | Generalized idiopathic epilepsy and epileptic syndromes, not intractable, without status epilepticus | 27 | 6.2 |
| G40.011 | Localization-related (focal) (partial) idiopathic epilepsy and epileptic syndromes with seizures of localized onset, intractable, with status epilepticus | 23 | 5.3 |
| R62.50 | Unspecified lack of expected normal physiological development in childhood | 15 | 3.4 |
| G40.812 | Lennox-Gastaut syndrome, not intractable, without status epilepticus | 10 | 2.3 |
| R56.9 | Unspecified convulsions | 10 | 2.3 |
| Q99.8 | Other specified chromosome abnormalities | 9 | 2.1 |
| Z15.89 | Genetic susceptibility to other disease | 8 | 1.8 |
| F84.0 | Autistic disorder | 7 | 1.6 |
| G40.109 | Localization-related (focal) (partial) symptomatic epilepsy and epileptic syndromes with simple partial seizures, not intractable, without status epilepticus | 7 | 1.6 |
